# Strong Effects of Population Density and Social Characteristics on Distribution of COVID-19 Infections in the United States

**DOI:** 10.1101/2020.05.08.20073239

**Authors:** Kumar B. Rajan, Klodian Dhana, Lisa L. Barnes, Neelum T. Aggarwal, Laura E. Evans, Elizabeth A. McAninch, Jennifer Weuve, Robert S. Wilson, Charles DeCarli, Denis A. Evans

## Abstract

**Background:** Coronavirus disease 2019 (COVID-19) has devastated global populations and has had a large impact in the United States. The objective of this manuscript is to study the relation of population demographics, social characteristics, and social distancing on the number of infections and deaths in the US.

**Methods:** Data came from publicly available sources. Social distancing was measured by the change in the rate of human encounters per km^2^ relative to the pre-COVID-19 national average. A smooth generalized additive model for counts of total infections and deaths at US county-level included population demographics, social characteristics, and social distancing.

**Results:** The model strongly predicted the geo-spatial variations in COVID-19 infections and deaths, 97.2% of variation in infections and 99.3% of variation in deaths from March 15, 2020. US counties with higher population density, poverty index, civilian population, and minorities, especially African Americans, had a higher rate of infections and deaths, and social distancing was associated with a slower rate of infections and deaths. The number of people infected was increasing; however, the rate of increase of new infections was showing signs of plateauing from the second week of April. Our model estimates that 1,865,580 US residents will test positive for infections and 117,246 fatalities by June 1, 2020. Importantly, our model suggests significant social differences in the infections and deaths across US communities. Areas with a larger African American population and a higher poverty index are expected to show higher rates of infections and deaths.

**Conclusion:** Preventive steps, including social distancing and community closures, have been a cornerstone in slowing the transmission and potentially reducing the spread of the disease. Crucial knowledge of the role of social characteristics in disease transmission is essential to understand and predict current and future disease distribution and plan additional preventive steps.

## INTRODUCTION

The COVID-19 is a global pandemic affecting 187 countries with over 3.84 million confirmed infections, 269,000 deaths, and a staggering fatality rate of 7.0% [1]. In the US alone, there are over 1.25 million infections, 76,000 deaths, and a fatality rate of 6.0% that has remained steady [2]. However, there are considerable variations in the COVID-19 infection and death rates across US communities over time. Hence, understanding the geospatial and temporal variation in COVID-19 infections and deaths requires serious and urgent attention. Many of these US communities show large variations in pre-existing chronic health conditions, population density, and socioeconomic status, as well as poorer access to essential protective resources and less ability to practice social distancing, all of which could lead to higher rates of infections and deaths.

The objective of this research manuscript is to develop a social transmission model to study geospatial temporal variation in infections and deaths across US counties. The social transmission model will utilize county-level population demographics. It will focus on population density, age distribution, and number of minorities [3-4], and social characteristics, such as poverty index [5-6], proportion of the county civilian employed labor force in white collar occupations [7], and social distancing using the rate of unique human encounters per km^2^ relative to the US national pre-COVID-19 baseline. This social transmission model will allow us to study the contribution of population demographics and social characteristics on the distribution of COVID-19 in US communities. Importantly, studying COVID-19 infections and deaths using our social transmission model will allow us to understand the predictors of current disease distribution across US communities, predict future distribution across US communities, and develop a national-level estimate. Most significantly, our social transmission model will help identify US communities that require the most resources to slow infection and death rates.

## METHODS

The data for this project comes from several compiled sources for testing data, daily infections, daily deaths, 2010 US census data, and data on social distancing. More details for each of these sources are provided below.

### Testing Statistics

The source for the total number of tests for COVID-19 came from the COVID tracking project [8] and the US Centers for Disease Control and Prevention (CDC) [9]. The COVID tracking project aggregates testing data by individual states and reports the number of people tested, including those tested in private labs. However, not all states report their figures, and this data should be considered a general indicator of testing output. The CDC reports on the specimens tested in the CDC labs and public health labs in 49 states, New York City, Puerto Rico, the United States Air Force, and 15 California counties. From these two sources, we would be able to obtain a general count of total US tests performed. However, the counts have up to seven days of lag when the specimens are accessioned, and testing is performed and summarized.

### Test Cases and Deaths

Several COVID-19 databases for infections and deaths have been made available for research purposes. We used county-level epidemiological data of confirmed cases and deaths, starting on March 1, 2020, which is available from Johns Hopkins University and updated with a daily time-series pattern [10]. Other epidemiological data, including situational reports from the World Health Organization (WHO) and Atlantic COVID-19 tracking projects, were also used to check the accuracy and reports. Data downloads from the sources were automatic, and a daily update was performed to obtain the most recent data.

### US Census Data

The US Census Bureau is the leading source of statistical information about people living in the US in the form of a decennial census. This census counts the entire US population every ten years (using a combination of long and short forms) and performs several other surveys [11]. The US census bureau collects several pieces of information from the population and presents the information using several hundred identified population and housing tables at the census block. The 2010 US census data has been downloaded, curated, and integrated with county-level infections and deaths.

### American Community Survey (ACS)

The ACS is an ongoing monthly survey sent to 3.5 million addresses to produce detailed population and housing estimates each year [12]. The ACS is designed to produce critical information on small geographic areas and release annual estimates for over 35,000 communities. The ACS collects several pieces of economic and community data that are relevant to this project. The ACS, performed through the census bureau, only started collecting more detailed data as of 2000. We used economic data from the 2008 ACS survey on the poverty index and the non-professional civilian population for each county.

### Social Distancing

According to the CDC and WHO, social distancing is currently the most effective way to slow the spread of COVID-19 through US communities. Unacast has developed a social distancing data program that consists of daily encounters, daily visitation, and daily non-essential visits compared to pre-COVID-19 and averaged for the US population [13]. We used the daily encounter rate since it provides the most appropriate information to study the change in human encounters per square km of residents in each US county.

### Statistical Analysis

Descriptive plots for infections and deaths summarized for all US counties provided information on cumulative infections and deaths. New daily infections and deaths were estimated based on the difference in cumulative infections and deaths between current and previous days. Similar characteristics were estimated for testing, hospitalization, and encounter rates across all US counties.

For our social transmission model, we used a smooth generalized additive model [14] with quasipoisson counts for total infections and deaths that included population density, poverty index, the proportion of non-Hispanic Whites, African Americans, and Hispanics, the proportion of females and non-professional civilians, age distributions (below 20, 20–40, 40–60, above 60), and social distancing for each US county. A county-level model was developed in several steps. The first step used the time since March 1, 2020, and latitudinal and longitudinal coordinates for counties, and explained 16% of the variation in the rate of confirmed infections. The addition of population demographics, social characteristics, and social distancing explained 98% of the variation in COVID-19 infections [15]. This model also included splines for time since March 1, 2020, population density, and latitude and longitude. In a separate model, we included testing characteristics and found the predictive models to be unstable due to extensive missing data at the county-level, underreporting, and severe lags. These additional variables were therefore excluded from our final model. A second social transmission model for COVID-19 deaths included all the variables of the infection model and the number of infections in each US county. All plots and statistical models were performed in Microsoft R Open Version 3.5.3 ×86_64-pc-linux-gnu (64-bit) and Intel MKL for parallel mathematical computing using 18 cores [16].

## RESULTS

According to the most recent estimate, 1.41 million US residents (or 4,225 residents per million) are infected with COVID-19. Of those infected, 85,500 US residents (or 254.2 residents per million) have died. The cumulative number of infections and deaths show a continual increase; however, the rate of growth in new COVID-19 infections (Figure 1A) shows a dramatic rise until April 8, 2020. Since then, the new infection rates show signs of plateauing with a slow downward trend. A similar increasing pattern in cumulative deaths was observed; however, the rate of deaths peaked on April 15, 2020, and was followed by a steady downward trend (Figure 1B). Thus, the figures for new COVID-19 infections and deaths may indicate that the peak of the US outbreak may have been reached.

**Figure 1.**
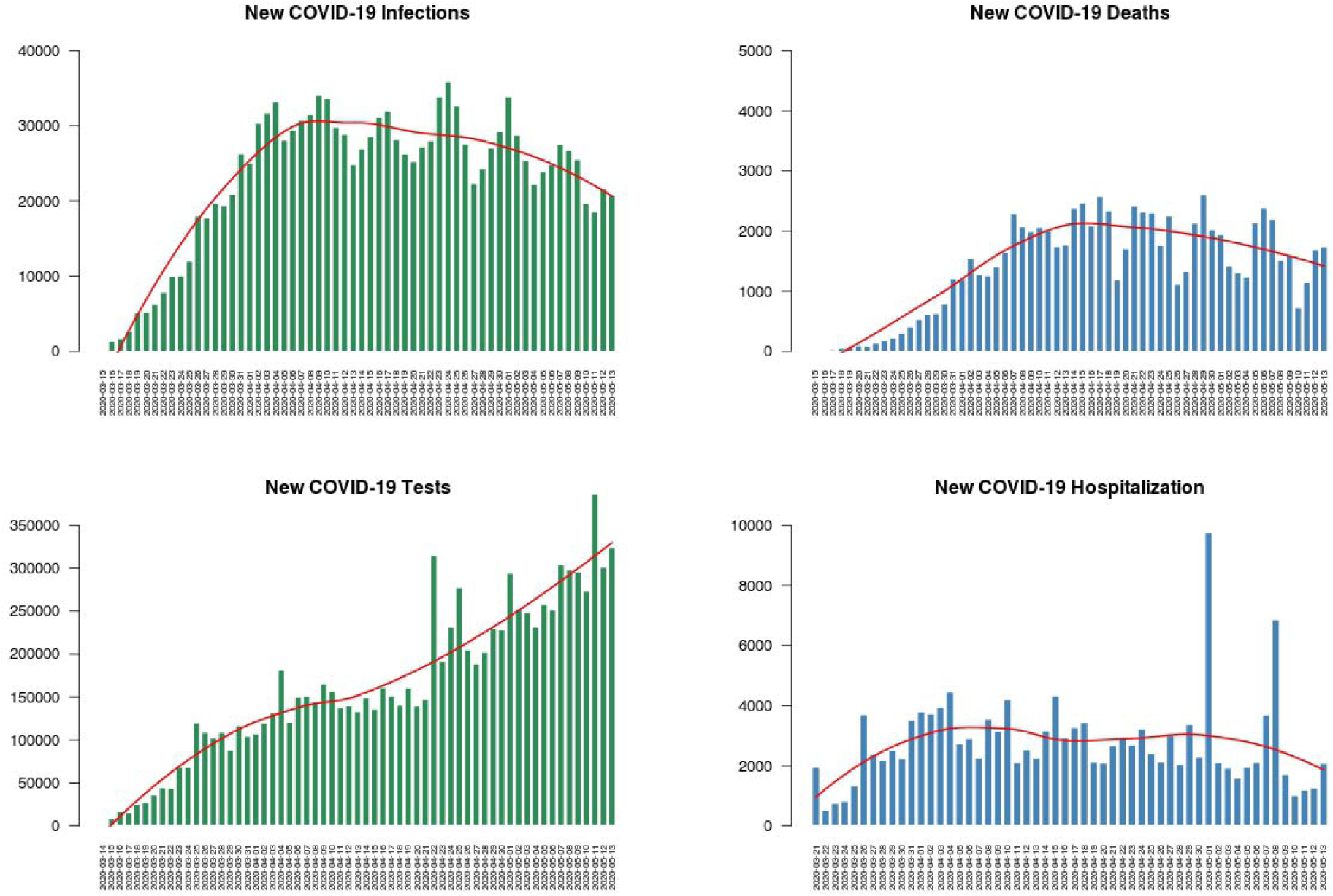
Daily New Covid-19 Infections, Deaths, Testing, and Hospitalization in the United States from March 15, 2020 The top left panel (green) shows the daily new Covid-19 infections and the top right panel shows the daily new Covid-19 deaths in the US. The bottom left panel (green) shows the daily new Covid-19 tests and the bottom right panel shows the daily new Covid-19 hospitalizations in the US. A loess smoother (red line) is fit to the daily new infections and deaths in the US.

Testing for COVID-19 has also steadily increased in the US, starting with 41,191 total tests on March 15, 2020, and rising to 9,965,819 on May 13, 2020. The daily increase in testing had risen to 105,423 by April 1, 2020, and to 218,465 by April 30, 2020. The highest daily increase in testing was 386,552 on May 11, 2020. Figure 1(C) shows the number of new daily tests in the US, which increased over time with 25,000–30,000 tests being performed daily as of May 1, 2020. In terms of US states, New York performed the most tests (over 2,000,000), with California testing half as often as New York, followed by Florida, Texas, and Washington.

According to the COVID tracking project, there were 149,347 hospitalizations for patients with COVID-19. The overall cumulative COVID-19-associated hospitalizations were 45.0 per 100,000 people, with the highest rates in people over the age of 65. Daily new hospitalizations in the US had been around 2,000 per day (Figure 1D). However, on May 1, new COVID-19 related hospitalizations showed a dramatic increase to 10,000, which then fell back to 2.000 new hospitalizations. Reporting on hospitalizations by the states had been inconsistent. New York had the most hospitalizations, followed by Connecticut, Massachusetts, and Florida.

We developed a social contagion model to predict the US distribution of COVID-19 infections and deaths from population demographics, social characteristics, and social distancing. The model for infections accounted for 97.2% of the variation in data, with 94.5% deviance. The model for deaths accounted for 99.3% of the variation in data, with 96.8% deviance across 3,364 US counties. The population demographics and social characteristics were also strongly associated with the rate of increase in confirmed COVID-19 infections. According to the social contagion model, we predict that 1,865,580 US residents will have confirmed COVID-19 infections, and 117,246 US residents will die by June 1, 2020. The actual (red line), estimated (blue line), and predicted (brown line) for COVID-19 infections are shown in Figure 2A and for COVID-19 deaths in Figure 2B.

**Figure 2.**
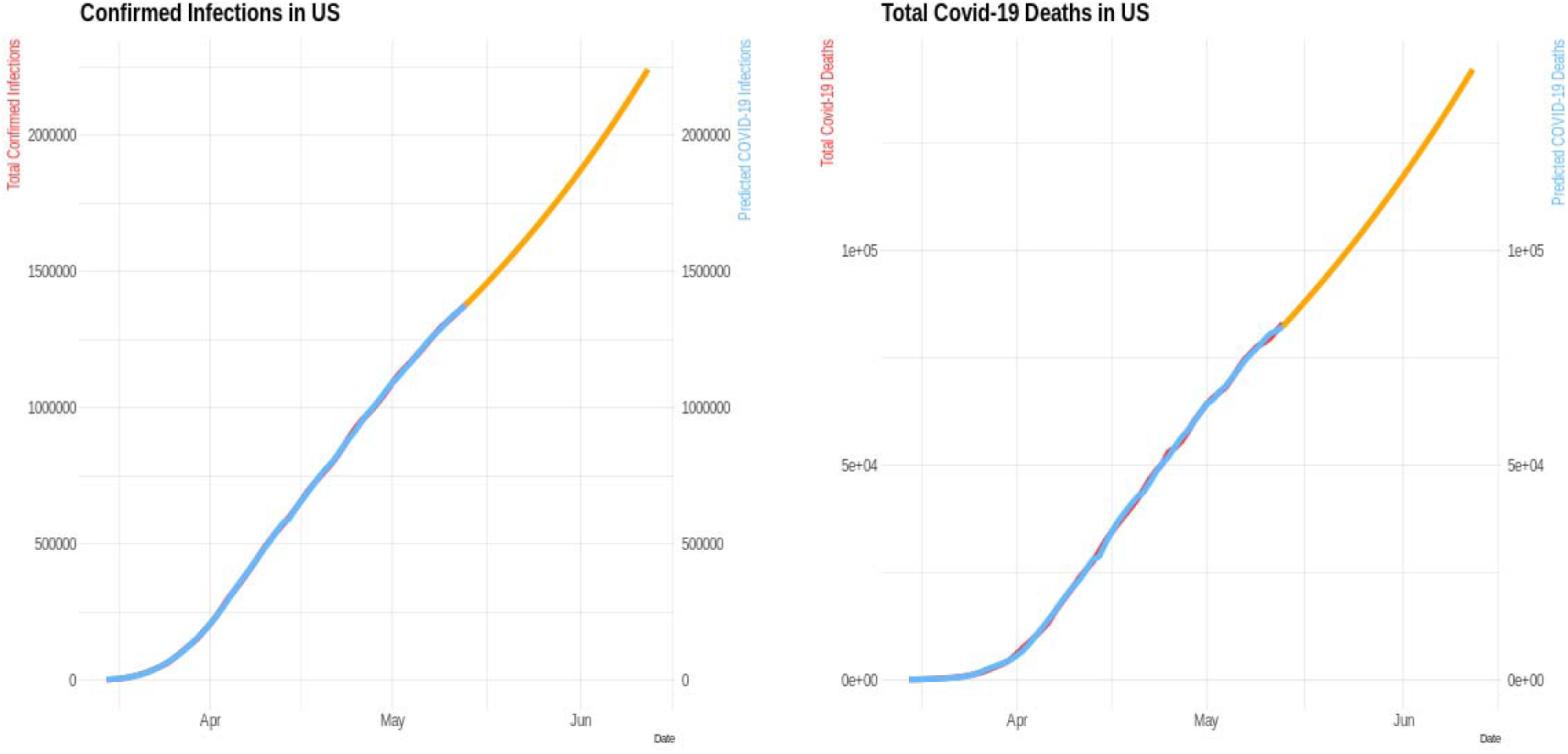
Cumulative Confirmed Infections and Deaths in the US from March 15, 2020 to June 1, 2020 with Actual and Estimated Values using the Social Transmission Model The left panel shows the confirmed infections and right panel the reported deaths in the US. In the figures, red line shows the actual confirmed Covid-19 infections and deaths, the blue line shows the estimated Covid-19 infections and deaths, and the brown line shows the estimated Covid-19 infections and deaths until June 1, 2020.

Geospatial variation in confirmed COVID-19 infections (Figure 3) and deaths (Figure 4) with population demographics and social characteristics from March 1 to June 1, 2020, is evident in time-lapsed US maps. Most of the eastern US and large counties in the western US show considerable increases in the number of cases. Many counties in the central US are not reporting COVID-19 cases due to incomplete data and lack of COVID-19 cases, but the exact reasons are harder to discern. Geospatial variations in confirmed COVID-19 deaths from March 1 to June 1, 2020, are starker, with much of the northeast, Midwest, southeast, southwest, and northwest showing higher death rates.

**Figure 3.**
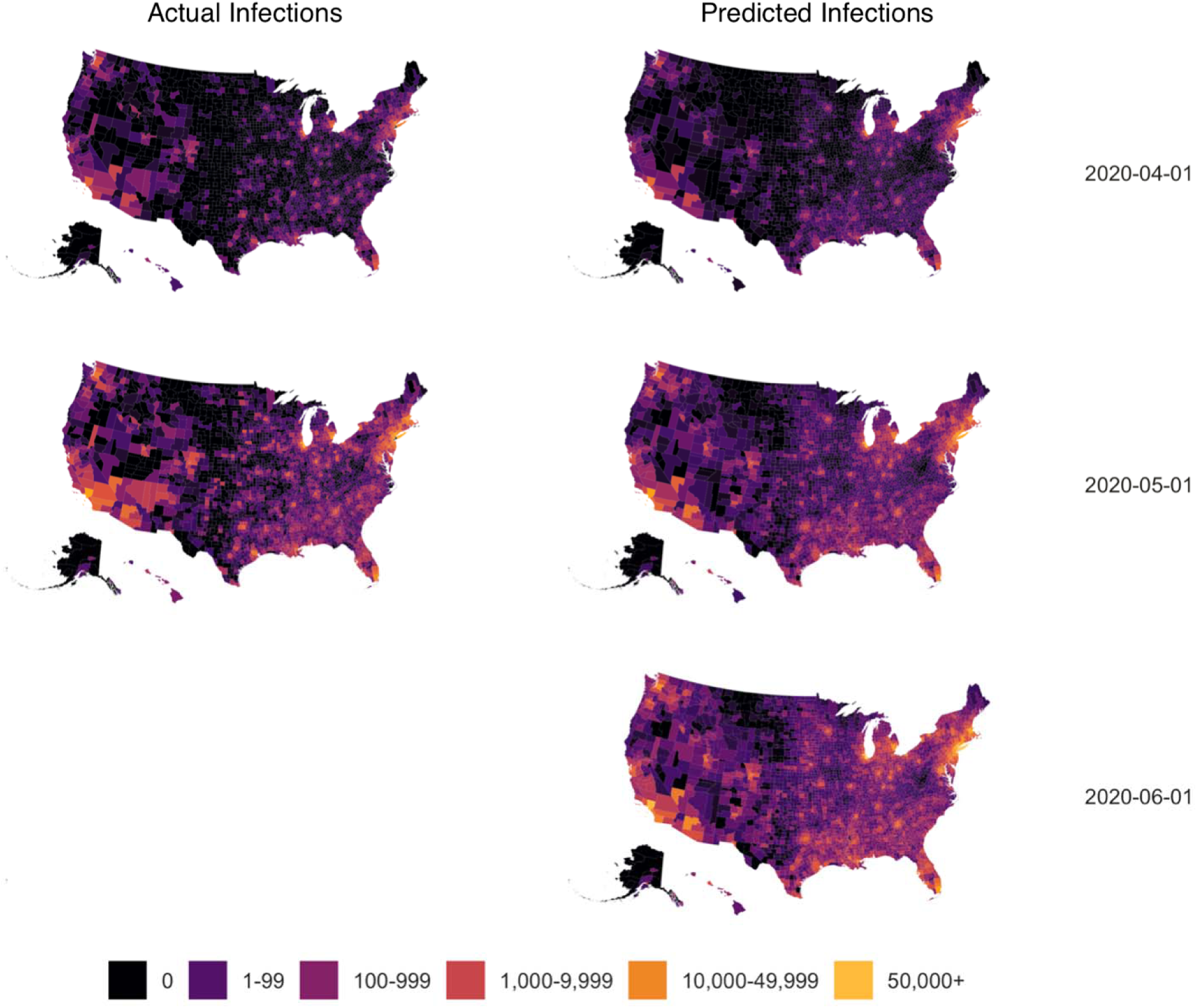
Distribution of Actual and Estimated Covid-19 Infections on Selected Dates from March 15 to June 1, 2020 in US Counties The left side shows the actual Covid-19 infections in US counties on April 1 and May 1, 2020. The right panel shows the estimated Covid-19 infections in US counties on April 1, May1, and June 1, 2020.

**Figure 4.**
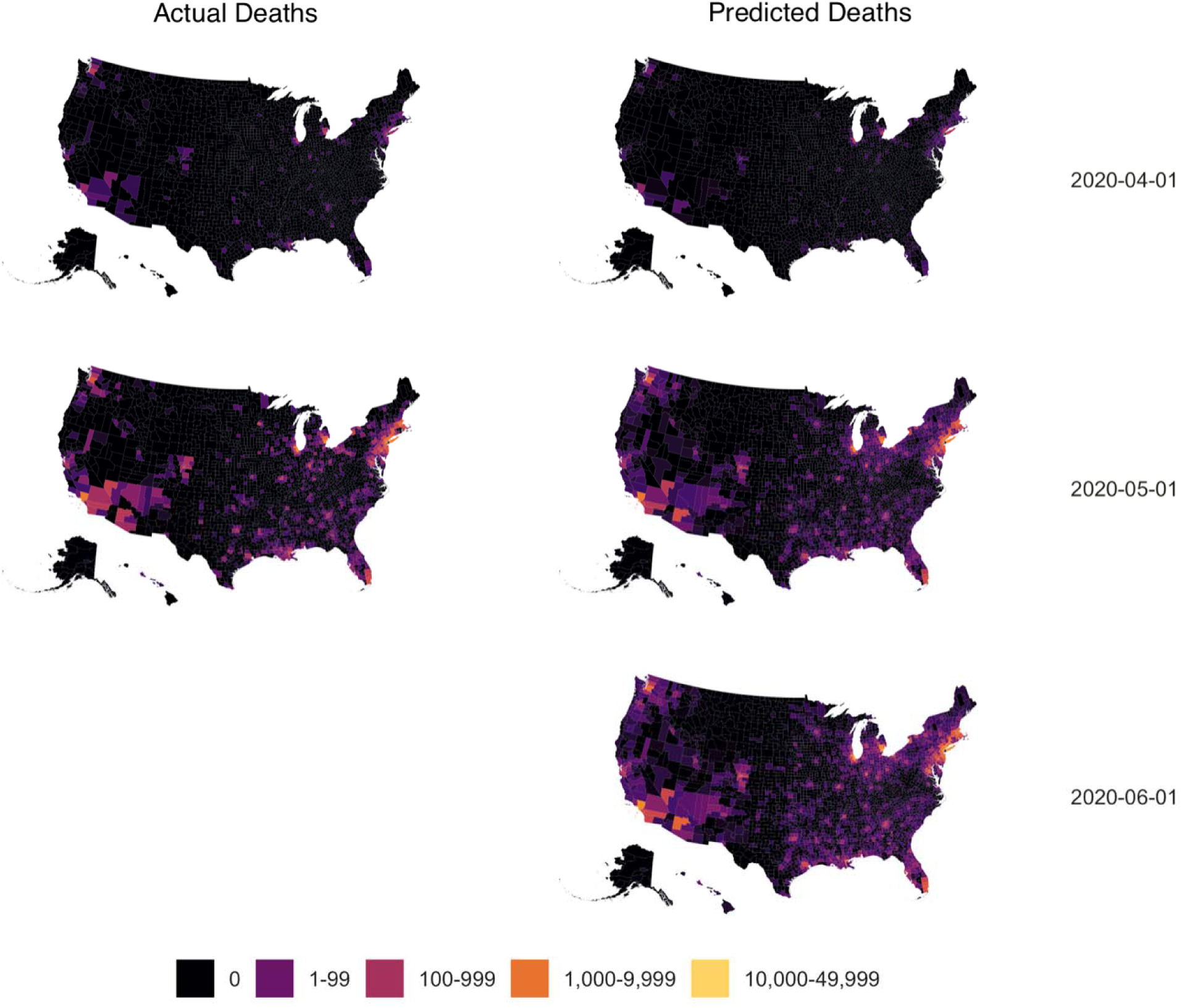
Distribution of Actual and Estimated Covid-19 Deaths on Selected Dates from March 15 to June 1, 2020 in US Counties The left side shows the actual Covid-19 deaths in US counties on April 1 and May 1, 2020. The right panel shows the estimated Covid-19 deaths in US counties on April 1, May1, and June 1, 2020.

In US counties with higher proportions of African Americans, the rate of COVID-19 infections increased by 5.4% for one percent increase African Americans (Figure 5), whereas the rate of increase was 2.3% in Whites and 4.9% in Hispanics. Additionally, in US counties with a higher poverty index, the rate of infections was 0.7% for a one-unit increase in the US census poverty index. In areas with a higher civilian employed labor force in white collar, the rate of infection was also higher. In US counties with a large young population (below 20) and a large older population (60–80), the rate of infections was higher. Rates of infection in counties with large younger and midlife populations (20-40 and 40-60) and a large oldest population (above 80) were lower. The rate of death increased by 0.8% for each unit increase in the poverty index, and 0.8% for one percent increase in African Americans (Figure 6), whereas the rates decreased by 0.8% for each percent increase in non-Hispanic Whites and 0.1% for each percent increase in Hispanics. Ages above 40 were associated with higher death rates, whereas ages below 40 were associated with lower death rates. Areas with higher number of adults over the age of 80 showed almost 6-fold higher deaths than those between 40-60. These findings suggest strong effects of population demographics and social characteristics on confirmed COVID-19 infections and deaths.

**Figure 5.**
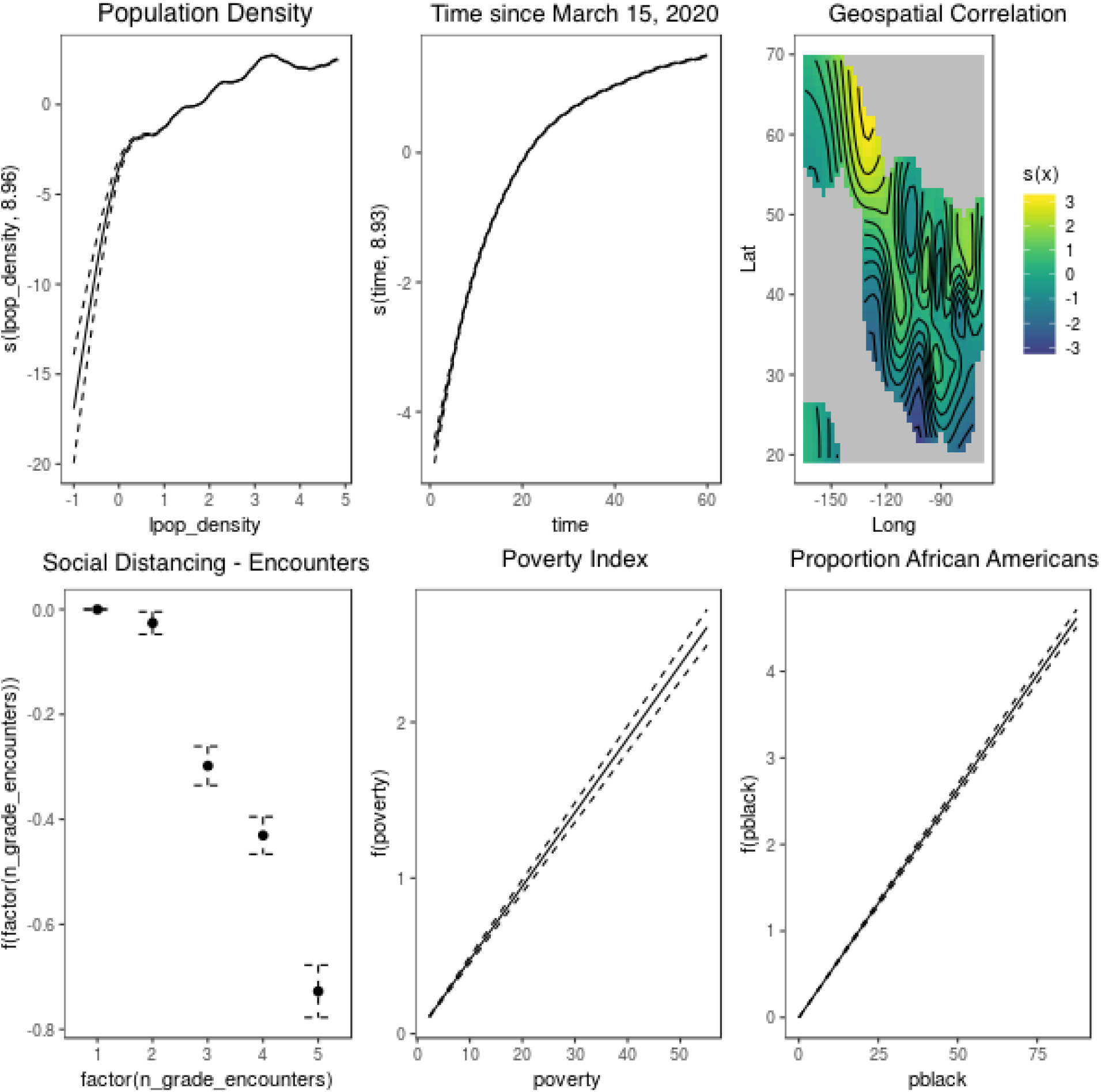
Relation of Population Density, Time, Geographical, Social Distancing, Poverty, and Percentage Blacks on Rate of Covid-19 Infections and Deaths in US Counties The rate of Covid-19 infections uses smooth terms for log10 transformed population
density, time since March 15, 2020, and geospatial characteristics, and additive terms for social distancing using encounter rates, poverty index, and proportion of non-Hispanic Blacks.

**Figure 6.**
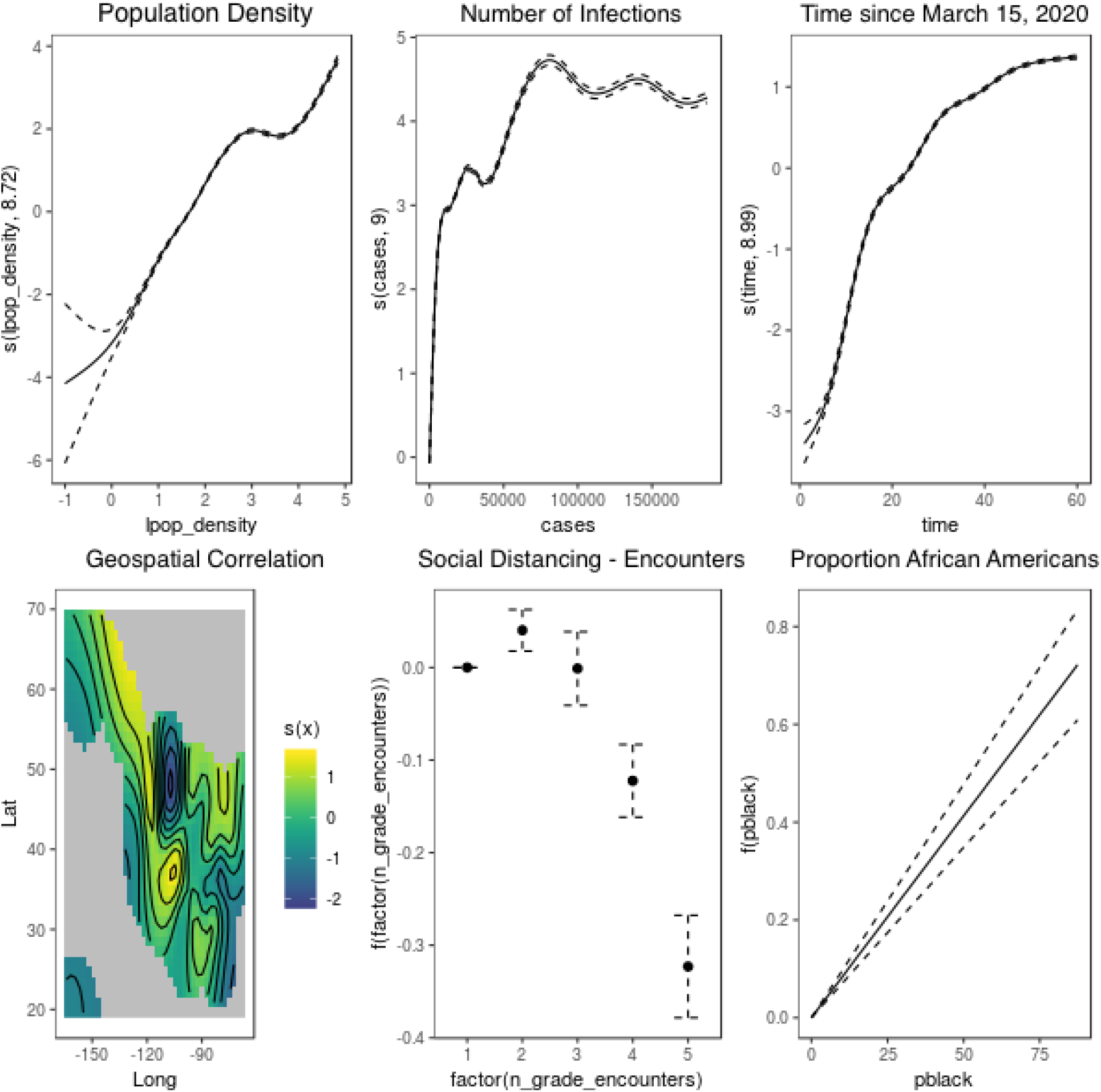
Relation of Population Density, Time, Geographical, Social Distancing, Poverty, and Percentage Blacks on Rate of Covid-19 Infections and Deaths in US Counties The rate of Covid-19 infections uses smooth terms for log10 transformed population density, time since March 15, 2020, and geospatial characteristics, and additive terms for social distancing using encounter rates, poverty index, and proportion of non-Hispanic Blacks.

Population density, social distancing, time, and geospatial variation were also associated with the number of confirmed COVID-19 infections (Figure 5) and deaths (Figure 6). In US counties with higher population density, the rate of increase of confirmed COVID-19 infections was exponentially higher. In 10% of US counties (N=311) with a population density higher than 500 residents per square mile, we observed 80% of total infections and deaths. As social distancing increased, the rate of COVID-19 infections and deaths was lower. The time trend showed a steep increase in the rate of infections from March 15 to about April 15, 2020, with the rate of infections leveling off and then slowing between April 15 and May 5, 2020.

Social distancing has a strong and consistent association with COVID-19 infections and deaths in US communities. Our social transmission model predicts that by June 1, 2020, the US will have 1,865,580 confirmed COVID-19 cases if the social distancing across all counties remains the same. If we see a 20% decline in social distancing, we project 46,433 additional COVID-19 infections and 66,764 additional infections if social distancing were to decrease by 30%. Social distancing also had a substantial influence on death. We project that by June 2020, the US will have 117,246 deaths attributed to COVID-19 if the social distancing across all counties remains the same. If we see a 20% decline in social distancing, we project 2,785 additional deaths, and 4,006 additional deaths if social distancing were to decrease by 30%.

## DISCUSSION

The social transmission model shows the high relevance and significance of population and social characteristics on COVID-19 infections and deaths in US. According to our model, US counties with high population density have high rates of COVID-19 infections and deaths, with rates of infections and deaths increasing exponentially. A small proportion (10%) of densely populated US counties account for 80% of all confirmed US infections and deaths. These densely populated communities had twice the number of non-Hispanic African Americans than communities that were less densely populated, similar to other health outcomes [3]. This is also reflected in our social contagion model, where rates of infections and deaths were dramatically higher in US counties with large numbers of non-Hispanic African Americans.

Community-level transmission was slower in communities with higher social distancing. As social distancing increased, the rate of increase of confirmed infections and deaths started to decline, suggesting that a substantial increase in confirmed infections and deaths may be attributable to a reduction in social distancing. High COVID-19 infections and deaths in densely populated areas were seen despite greater social distancing. Also, of significance is that communities with a high poverty index and related social characteristics, in general, had lower social distancing compared to geographical areas with a low poverty index and similar social distancing. If social distancing restrictions were to be reduced in these densely populated lower socioeconomic areas, we might see a higher number of confirmed infections and deaths in these communities. If social distancing can be improved in densely populated areas with high poverty indices, we might see substantial reductions in confirmed infections and deaths. However, areas with a higher proportion of non-Hispanic African Americans showed a significantly higher rate of infection and deaths, and these effects were larger than the social distancing effects, which, in general, were more protective in areas with a higher proportion of non-Hispanic Whites, a similar phenomenon reported previously [5]. Our findings suggest the strong effects of race/ethnicity on infections and deaths in US communities.

Age distribution plays a significant role in the rate of increase in COVID-19 confirmed infections and deaths. It is noteworthy that areas with high infections and deaths also had larger numbers of younger residents, below 20 years old, and a larger number of residents between the ages of 60 and 80. In areas with a higher number of middle-aged adults, 20-40 and 40–60 years old had a lower rate of infection, suggesting that more social distancing is being maintained in the middle-aged groups than in the younger age groups. Areas with higher number of adults over the age of 80 showed dramatically higher mortality rates, since older individuals a high susceptibility for adverse outcomes due to the greater amount of chronic health conditions associated with age.

The rate of change in the number of infections and deaths increased exponentially in late March and early to mid-April. However, the rate of new infections has stabilized over time, reaching a plateau, where it remains steady. The rates of new infection and death across the US communities have started to decline, due to social distancing. However, the evidence for a continued decline over 14 days, as suggested by the US government, has yet to be observed in the most densely populated areas. The stable rate of new infections and lack of data on COVID-19 deaths from many counties are troublesome. Even a phased re-opening in densely populated US communities may cause a large increase in infections and deaths unless more precautions and preventive measures are put in place.

The social transmission model provides a framework for incorporating population demographics and social characteristics, in addition to temporal and geospatial patterns, as predictors of COVID-19 infections and deaths in US communities. Even if testing were to be dramatically increased, this approach alone does not address the highly infectious nature of COVID-19. The major role of population demographics and social characteristics on infection rates may be mitigated by social distancing. Focusing our preventive efforts on population centers with a higher number of non-Hispanic African Americans, and areas of high poverty, and providing the necessary resources to increase social distancing and testing in those areas might offer a better chance of reducing the spread of COVID-19 and deaths associated with COVID-19 across US communities.

## Data Availability

All data have been collected from publicly available sources and will be made available upon request.

